# Common genetic risk variants identified in the SPARK cohort implicate *DDHD2* as a novel autism risk gene

**DOI:** 10.1101/2020.01.13.20017319

**Authors:** Nana Matoba, Dan Liang, Huaigu Sun, Nil Aygün, Jessica C. McAfee, Jessica E. Davis, Laura M. Raffield, Huijun Qian, Joseph Piven, Yun Li, Sriam Kosuri, Hyejung Won, Jason L. Stein

**Affiliations:** Department of Genetics, University of North Carolina at Chapel Hill, North Carolina, USA; UNC Neuroscience Center, University of North Carolina at Chapel Hill, North Carolina, USA; Department of Chemistry and Biochemistry, University of California, Los Angeles, Los Angeles, California, USA; UCLA-DOE Institute for Genomics and Proteomics, University of California, Los Angeles, Los Angeles, California, USA; Molecular Biology Institute, University of California, Los Angeles, Los Angeles, California, USA; Quantitative and Computational Biology Institute, University of California, Los Angeles, Los Angeles, California, USA; Eli and Edythe Broad Center of Regenerative Medicine and Stem Cell Research, University of California, Los Angeles, Los Angeles, California, USA; Jonsson Comprehensive Cancer Center, University of California, Los Angeles, Los Angeles, California, USA; Department of Statistics and Operations Research, University of North Carolina at Chapel Hill, North Carolina, USA; Department of Psychiatry and the Carolina Institute for Developmental Disabilities, University of North Carolina at Chapel Hill, North Carolina, USA; Department of Biostatistics, University of North Carolina at Chapel Hill, North Carolina, USA; Department of Computer Science, University of North Carolina at Chapel Hill, North Carolina, USA

**Keywords:** Autism spectrum disorder, GWAS, common variant, MPRA, DDHD2

## Abstract

**Background:** Autism spectrum disorder (ASD) is a highly heritable neurodevelopmental disorder. Large genetically informative cohorts of individuals with ASD have led to the identification of three common genome-wide significant (GWS) risk loci to date. However, many more common genetic variants are expected to contribute to ASD risk given the high heritability. Here, we performed a genome-wide association study (GWAS) using the Simons Foundation Powering Autism Research for Knowledge (SPARK) dataset to identify additional common genetic risk factors and molecular mechanisms underlying risk for ASD.

**Methods:** We performed an association study on 6,222 case-pseudocontrol pairs from SPARK and meta-analyzed with a previous GWAS. We integrated gene regulatory annotations to map non-coding risk variants to their regulated genes. Further, we performed a massively parallel reporter assay (MPRA) to identify causal variant(s) within a novel risk locus.

**Results:** We identified one novel GWS locus from the SPARK GWAS. The meta-analysis identified four significant loci, including an additional novel locus. We observed significant enrichment of ASD heritability within regulatory regions of the developing cortex, indicating that disruption of gene regulation during neurodevelopment is critical for ASD risk. The MPRA identified one variant at the novel locus with strong impacts on gene regulation (rs7001340), and expression quantitative trait loci data demonstrated an association between the risk allele and decreased expression of *DDHD2* (DDHD domain containing 2) in both adult and pre-natal brains.

**Conclusions:** By integrating genetic association data with multi-omic gene regulatory annotations and experimental validation, we fine-mapped a causal risk variant and demonstrated that *DDHD2* is a novel gene associated with ASD risk.

## Introduction

Autism spectrum disorder (ASD) is a common neurodevelopmental disorder characterized by characteristic social deficits as well as ritualistic behaviors (1). Because ASD is highly heritable (∼50-80%) (2–6), a number of studies have been conducted to identify both rare and common genetic variants contributing to risk for ASD. While previous studies have successfully identified rare *de novo* presumed loss of function mutations leading to risk for ASD (7–13), these *de novo* variants do not explain the large heritability and therefore are missing an important component of ASD risk.

To identify common inherited genetic risk factors, genome-wide association studies (GWAS) have now accumulated over 18,000 individuals with ASD and have begun discovering genome-wide significant (GWS) loci that explain some of the inherited risks for ASD (14). The three GWS ASD susceptibility loci discovered previously explain in total only 0.13% of the liability for autism risk, whereas all common variants are estimated to explain 11.8% of liability (14). Therefore, there are more common risk variants to be discovered, which requires larger sample sizes to provide sufficient power to detect risk variants of small effect (15–17). The newly established genetic cohort, SPARK (Simons Foundation Powering Autism Research for Knowledge) (https://sparkforautism.org/) is planning to collect and analyze data from 50,000 individuals with ASD (18). SPARK has recently released genotype data for over 8,000 families or singletons with ASD, which we utilize here to increase the power of ASD GWAS.

Once we identify GWS loci, the critical next step is to understand their biological impact. This is especially challenging because most GWAS identified loci for neurodevelopmental disorders and other traits are located in poorly annotated non-coding regions with presumed gene regulatory function (19). In addition, most loci are comprised of multiple single nucleotide polymorphisms (SNPs) that are often inherited together, which makes it difficult to identify the true causal variant(s) and their regulatory effects (20,21). To overcome these problems, various experimental validation tools have been developed (22–24). One of these tools, a massively parallel reporter assay (MPRA), simultaneously evaluates allelic effects on enhancer activity for many variants. In this assay, exogenous DNA constructs, harboring risk and protective alleles at an associated variant, drive the expression of a barcoded transcript. Differences in barcode counts between the risk and protective alleles indicate the regulatory function of that variant (23,24). This assay thus demonstrates the regulatory potential of individual SNPs and provides evidence of the causal variants within an associated locus.

Though fine-mapping approaches can suggest causal variants at a locus, they cannot identify target genes affected by those variants. Several approaches are designed to link variants to genes they regulate including expression quantitative trait loci (eQTL) (25–27) as well as chromatin interaction (via Hi-C) assays (28–30). Recently, we developed Hi-C coupled MAGMA (H-MAGMA) which predicts genes associated with the target phenotype by integrating long-range chromatin interaction with GWAS summary statistics (31). Together with existing eQTL resources in the adult and fetal cortex (32,33), it is possible to link variants associated with risk for ASD to target genes and functional pathways.

In this study, we increase the sample size of existing ASD GWAS by adding 6,222 cases-pseudocontrol pairs from the genetically diverse SPARK project. Our analysis identified five loci associated with risk for ASD including two novel loci. For one novel locus identified, we used an MPRA to identify the causal variant within the locus. Further, we integrated multi-level functional genomic data obtained from the developing brain, including eQTLs, chromatin interactions, and regulatory elements, to identify *DDHD2* as a candidate gene involved in ASD etiology at the MPRA-validated locus.

## Methods and Materials

This study (analysis of this publicly available dataset) was reviewed by the Office of Human Research Ethics at UNC, which has determined that this study does not constitute human subjects research as defined under federal regulations [45 CFR 46.102 (d or f) and 21 CFR 56.102(c)(e)(l)] and does not require IRB approval.

### SPARK dataset

SPARK participants who received any of the following diagnoses: autism spectrum disorder [ASD], Asperger syndrome, autism/autistic disorder and pervasive developmental disorder-not otherwise specified (PDD-NOS) were recruited. The samples were enriched for affected individuals whose parents were also available to participate. Participants registered for SPARK online at www.SPARKforAutism.org or at 25 clinical sites across the country by completing questionnaires on medical history and social communication as described here: https://www.sfari.org/spark-phenotypic-measures/.

In the current study, participants were drawn from the SPARK 27K release (20190501 ver.) through SFARIBase (https://www.sfari.org/resource/sfari-base/), which included 27,290 individuals (who were genotyped with a SNP array and/or whole-exome sequencing [WES]) with phenotype information such as sex, diagnosis, and cognitive impairment. The data included probands and their family members if applicable (e.g. 3,192 quads (2,798 families with unaffected siblings, 394 with multiple affected siblings), 2,486 trios, and 2,448 duos) (Supplementary Figure S1). Twenty families in this release overlapped with the Simon’s Variations in Individuals Project (SVIP) cohort and were subsequently removed for the genome-wide association analysis (Supplementary Figure S2) since the SVIP cohort has targeted probands with 16p11.2 deletions. We also obtained whole-exome sequencing (WES) data to estimate the imputation accuracy. Details on genotyping and whole-exome sequencing data, and pre-imputation quality control are provided in Supplementary Methods.

### Genotype phasing and imputation

Phasing was performed using EAGLE v2.4.1 (34) (https://data.broadinstitute.org/alkesgroup/Eagle/) within SPARK samples. Before making pseudocontrols, we removed two individuals, one each from two pairs of monozygotic twins with Identity-By-Descent (PI_HAT)>0.9, by selecting the individual with lower call rates. We then defined pseudocontrols by PLINK 1.9 (35) (www.cog-genomics.org/plink/1.9/) for trios by selecting the alleles not inherited from the parents to the case (36). We re-phased all SPARK samples that passed our QC measures with pseudocontrols. Imputation was performed on the Michigan imputation server (37) (https://imputationserver.sph.umich.edu/index.html). Since SPARK participants are genetically diverse, we imputed genotypes using the Trans-Omics for Precision Medicine (TOPMed) Freeze 5b (https://www.nhlbiwgs.org/) reference panel which consists of 125,568 haplotypes from multiple ancestries (38,39). Imputation accuracy relative to WES was assessed using a similar approach to previous work (40) (Supplementary Figure S4) as described in Supplementary Methods.

### Genome-wide association analysis and Meta-analysis with iPSYCH-PGC study

We tested association within the SPARK all case-pseudocontrol pairs (full dataset; Supplementary Table S1) using PLINK2 generalized linear model (--glm) for SNPs with MAF ≥ 0.01 and imputation quality score from minimac4 (R2) > 0.5 (Supplementary Figure S4). In this model, we did not include any covariates since cases and pseudocontrols are matched on environmental variables and genetic ancestry. We performed secondary GWAS analyses by subsetting to only specific ancestry groups. We called ancestry using multidimensional scaling (MDS) analysis with 988 HapMap3 individuals and one random case from each trio (Supplementary Figure S3, Supplementary Table S2). Ancestry of individuals from SPARK was called as European, African or East Asian ancestries if they were within 5 standard deviations of defined HapMap3 population (CEU/TSI; YRI/LWK; or CHB/CHD/JPT, respectively) centroids in MDS dimensions 1 and 2. Population-specific GWASs were carried out using the same association model as described above for the SPARK all ancestries dataset. Meta-analyses with iPSYCH-PGC study (14) were performed by METAL (release 2018-08-28) (41). Additional information for iPSYCH-PGC summary statistics is provided in Supplementary Methods.

### Investigation of pleiotropic effects for ASD loci

The pleiotropic effects of identified loci were investigated for phenotypes available in the NHGRI/EBI GWAS catalog (downloaded October 22, 2019) (42) (Supplementary Methods).

### Linkage Disequilibrium Score regression analysis

LD SCore regression (LDSC) (v1.0.0) (43,44) was used to estimate genome-wide SNP based heritability, heritability enrichment of tissue/cell-type specific epigenetic states, and genetic correlation across phenotypes for GWAS meta-analysis result (Supplementary Methods). Prior to the analyses, we filtered SNPs to those found in HapMap3 and converted to LDSC input files (.sumstats.gz) using munge_sumstats.py. The pre-computed LD scores for Europeans were obtained from https://data.broadinstitute.org/alkesgroup/LDSCORE/eur_w_ld_chr.tar.bz2. For all LDSC analyses, we used individuals from European ancestry as described in the “Genome-wide association analysis (GWAS)” section above.

### Estimating polygenic risk score

Polygenic risk scores (PRSs) were calculated based on the iPSYCH-PGC study (14) using PRSice-2 (45) (https://www.prsice.info/). Details on generation of PRS, sex-stratified and family-type PRS, and parental origin PRS analyses are provided in Supplementary Methods.

### H-MAGMA

SNP to Ensembl gene annotation was carried out by Hi-C coupled MAGMA (H-MAGMA) (https://github.com/thewonlab/H-MAGMA) by leveraging chromatin-interaction generated from fetal brain Hi-C (46) as previously described (31). Details on H-MAGMA and functional analyses of H-MAGMA genes are provided in Supplementary Methods.

### Construction of a Massively Parallel Reporter Assay (MPRA) Library

Because the novel SPARK associated locus (chr8:38.19M - chr8:38.45M) was also detected in a previous schizophrenia GWAS which is better powered, we obtained credible set SNPs for the locus based on schizophrenia GWAS results (47) (see Supplementary Methods). Ninety-eight credible set SNPs were detected in this locus. We obtained 150bp sequences that flank each credible set SNP with the SNP at the center (74bp + 75bp). Because each SNP has risk and protective alleles, this resulted in 196 total alleles to be tested. We seeded HEK293 cells (ATCC® CRL-11268(tm)) in 6 wells (total 6 replicates) to be 70-90% confluent at transfection. We used lipofectamine 2000 (Invitrogen cat#11668) with our final MPRA library following the manufacturer’s instructions. Additional information for construction of MPRA library is available in Supplementary Methods. MPRA data was analyzed by an mpra package in R (48,49) (https://github.com/hansenlab/mpra) with more details in Supplementary Methods.

### Functional annotation of rs7001340 locus with multi-omic datasets

To investigate the target genes affected by allelic variation at rs7001340, we used two expression quantitative loci (eQTL) datasets derived from fetal cortical brain tissue and adult dorsolateral prefrontal cortex (32). We also used chromatin accessibility profiles from primary human neural progenitor cells and their differentiated neuronal progeny (unpublished data from Stein lab). Further information is provided in Supplementary Methods.

## Results

### GWAS in SPARK dataset identified a new locus associated with ASD risk

We obtained genotype and clinical diagnosis of ASD via self- or parent-report from 27,290 individuals who participated in the SPARK project (18). The majority of data comprised families, including those where both parents and multiple children were genotyped (quads; *N*=3,192 families), where both parents and one child were genotyped (trios; *N=*2,486 families), or where one parent and one child were genotyped (duos; *N*=2,448 families) (Supplementary Figure S1). Only 68 individuals were ascertained without family members (singletons). After genotyping quality control (Supplementary Methods), 375,918 variants from 26,883 individuals were retained. Because the SPARK dataset did not genotype unrelated controls, we created pseudocontrols from the alleles not transmitted from parents to probands (36). Case-pseudocontrol design requires genotyping of both parents, so singletons and duos were excluded from the analysis. Due to the diverse ancestry in the cohort (Supplementary Figure S2, Supplementary Table S1), genotypes of all individuals including pseudocontrols were imputed to a diverse reference panel (TOPMed Freeze 5b reference panel consisting of 125,568 haplotypes). After imputation quality control (Methods; Supplementary Figure S3), 90,051,896 autosomal SNPs were tested for association in 6,222 case-pseudocontrol pairs (SPARK full dataset) consisting of 4,956 males and 1,266 females from multiple ancestries including European (*N* = 4,535), African (*N* = 37), East Asian (*N* = 83) and other ancestries/admixed individuals (*N* = 1,567) (Supplementary Figure S1, Supplementary Table S2). We observed no inflation of test statistics (λ_GC_ = 1.00) (Supplementary Figure S3), indicating population stratification was well-controlled when using this case-pseudocontrol design. We identified two SNPs at one locus (index SNP: rs60527016-C; OR = 0.84, *P* = 4.70×10^−8^) at genome-wide significance (*P* < 5.0 × 10^−8^) (Figure 1A, Table 1), which were supported by the previous largest ASD GWAS (14) (OR = 0.95, *P* = 0.0047) derived from the PGC and iPSYCH cohorts (Supplementary Figure S4).

**Table 1.**
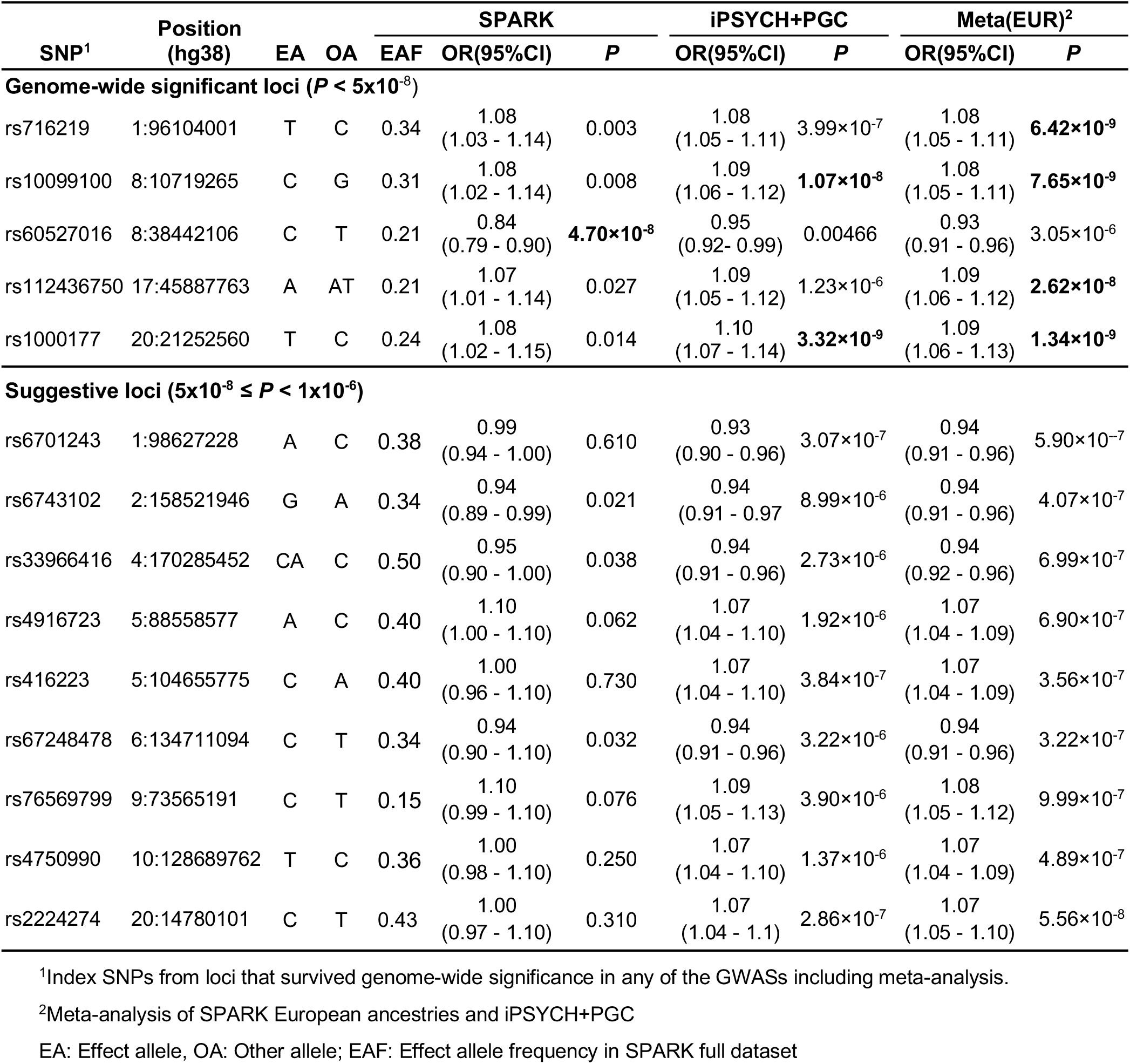
Genome-wide significant loci associated with ASD risk. Genome-wide significant and suggestive loci in any of the GWAS analyses and meta-analysis of SPARK European ancestries and iPSYCH+PGC participants are shown.

**Figure 1.**
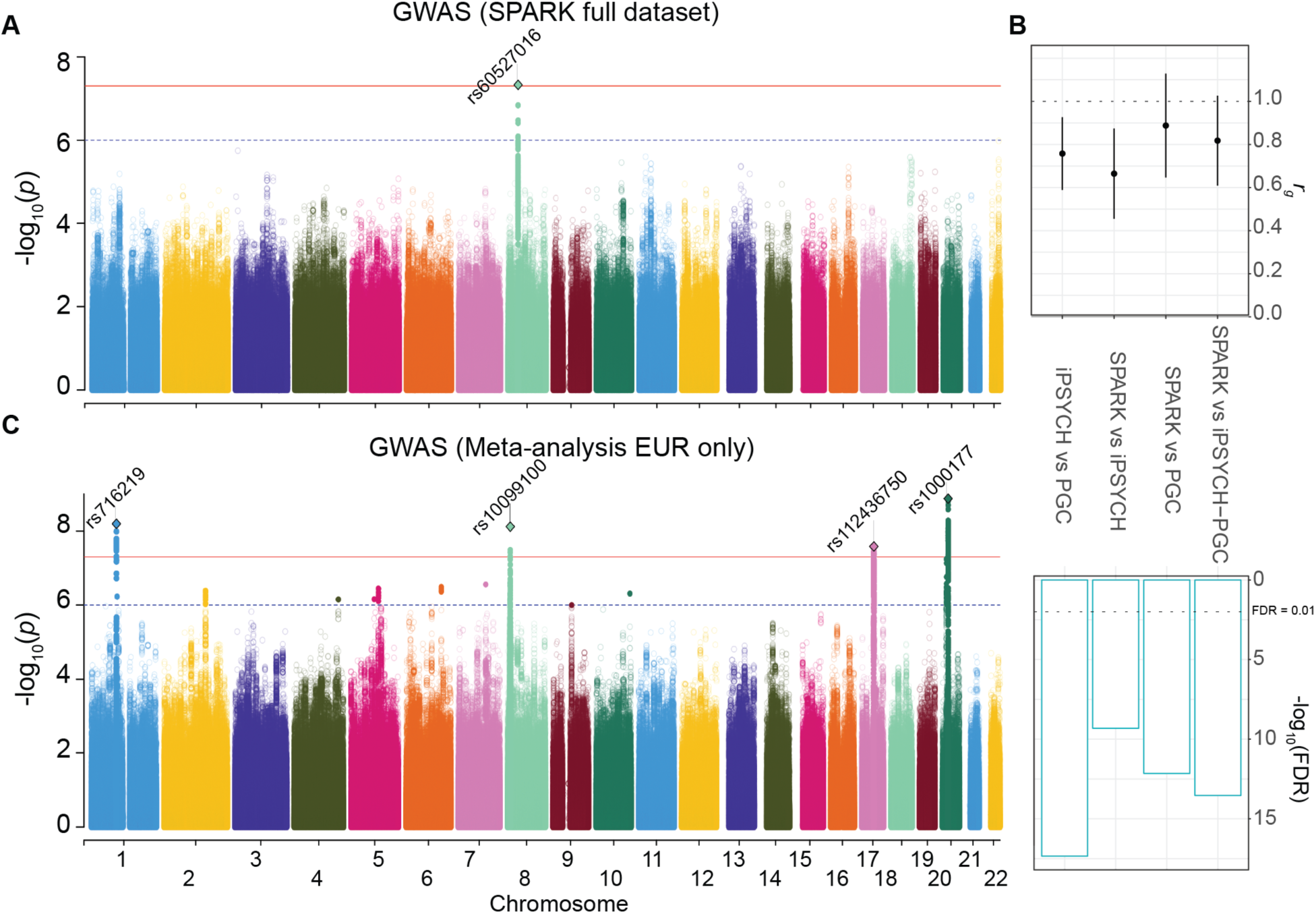
Genome-wide association of ASD in the SPARK dataset. (**A**) GWAS result from SPARK full dataset (N_case+pseudocontrol_ = 12,444). (**B**) Genetic correlations across ASD GWAS. From left to right, iPSYCH versus PGC (50), SPARK EUR versus iPSYCH, SPARK EUR versus PGC and SPARK EUR versus iPSYCH-PGC study (14). (**C**) GWAS results from the meta-analysis (SPARK European population and iPSYCH-PGC, N_max_case+control_ = 55,420). For Manhattan plots (**A, C**), the x-axes indicate the chromosomal position and y-axes indicate the significance of associations. The blue and red lines denote the significance threshold at suggestive (P < 1×10^−6^) and significant (P<5×10^−8^) levels. SNPs at with P < 1×10^−6^ are shown as a filled circle. Rs number indicates index SNPs from independent loci (1 MB apart from each other) at P < 1×10^−8^). Index SNPs at P < 5×10^−8^ are shown as diamonds.

### Replication of genetic risk factors for ASD

Given the phenotypic heterogeneity of ASD and potential technical differences such as genotyping platforms or data processing, we assessed the replication of genetic risk factors across cohorts by comparing previous major ASD studies including PGC and iPSYCH cohort (14,50) with the SPARK dataset subset to individuals of European descent (EUR) (Figure 1B). Although each study included multiple ASD subtypes including ASD from DSM5, Asperger’s, autism/autistic disorder, and Pervasive Developmental Disorder - Not otherwise specified (PDD-NOS) from DSM IV, and approaches differed across these samples from requiring community diagnosis to best-estimate diagnosis based on standardized assessment, we obtained high genetic correlations between the SPARK EUR dataset and the largest iPSYCH-PGC GWAS (*r_g_*= 0.82; *P* = 5.27 × 10^−14^), suggesting the genetic risk factors for autism are largely shared among different ASD GWAS and are generalizable despite differences in diagnostic criteria and batch effects. We next performed meta-analysis with SPARK EUR samples and iPSYCH-PGC samples (*N_case_* = 18,381 and *N_control_* = 27,969) to maximize power. The meta-analysis identified four additional loci associated with risk for ASD (Supplementary Figure S5-8). These included three previously reported loci (14) and one novel locus on chromosome 17, where a gene-based test from the iPSYCH-PGC study has previously shown association with risk for ASD (14) (Figure 1C, Table 1, Supplementary Figure S7). This novel locus was also reported to be associated with more than 60 phenotypes including neuroticism (51–55), educational attainment (56) and intracranial volume (57) (index SNPs *r*^*2*^> 0.8 in 1KG EUR) (Supplementary Table S3) indicating highly pleiotropic effects at this locus. The SNP based heritability in SPARK EUR samples was estimated (*h*^*2*^_*G*_) to be 0.117 (s.e. = 0.0082) for population prevalence of 0.012 (14,58) which was comparable with the previous report (*h*^*2*^_*G*_ = 0.118; s.e. = 0.010) (14).

The generalization of effects across ancestries for the five index SNPs identified (Table 1) was examined (Supplementary Figure S2, S3, and Supplementary Table S4). The association results from the cross-ancestry dataset were mainly driven by the European population, as expected given the larger sampling from this population. We found that some regions showed population-differentiated allele frequencies. For example, rs10099100 was more common in European and African populations (MAF = 0.33, 0.39 from tested samples, respectively) than in East Asians (MAF = 0.02 from tested samples), necessitating a further investigation of genetic risk factors for ASD in populations of diverse ancestry (59,60).

The generalization of genetic effects on risk for ASD was also confirmed by polygenic risk scores (PRSs) derived from the iPSYCH-PGC GWAS that showed higher scores in SPARK EUR cases (*N* = 4,097) compared to pseudocontrols (*N* = 4,097) (*P* = 1.61 × 10^−19^) (Figure 2A).

**Figure 2.**
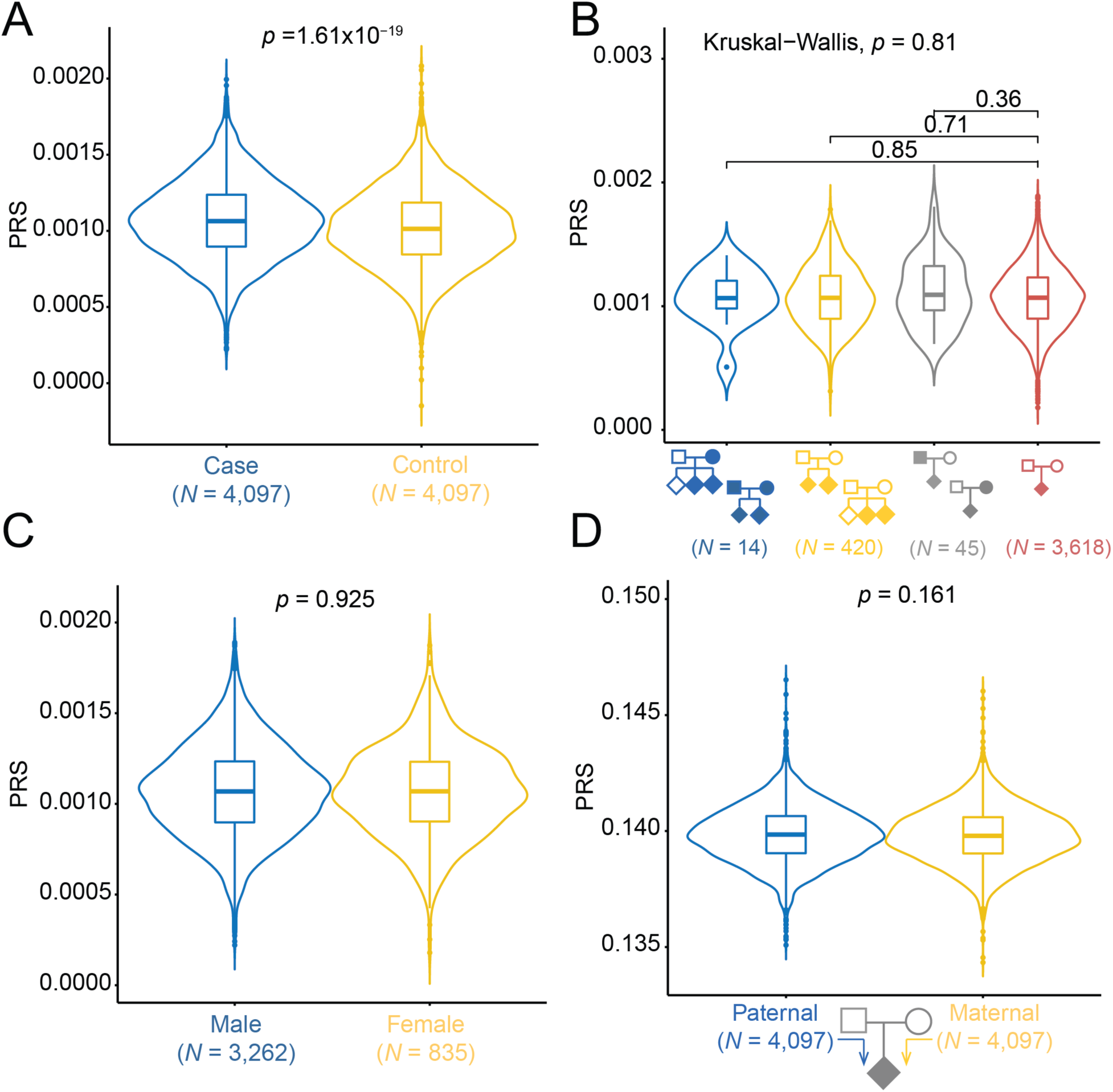
Comparison of polygenic risk scores between subgroups. **(A)** Common variant risk burden is higher in cases compared to pseudocontrols. **(B)** Comparison of PRS across family types (from left to right, families with multiple affected children with affected parent(s), multiple affected children with unaffected parents, one affected child with affected parent(s), and one affected child with unaffected parents) shows no evidence for a higher common variant burden in multiplex families. **(C)** Comparison of PRS between male and female probands shows no evidence of enrichment of common variants impacting risk for ASD in females. (**D**) There is no evidence for a difference in the transmission of common variant risk burden from mother versus father.

### Investigation of common variant burden impacting risk for ASD

We next used PRSs to compare common variant risk burden among family types, sex, and parent of origin (Figure 2B-D). Because ASD families with multiple affected siblings were shown to have different segregation patterns compared with simplex families that have a higher burden of de *novo* mutations (12,61,62), we compared the distribution of PRSs across four family types (Figure 2B, Supplementary Table S1). Our results showed no evidence for a difference in common variant burden impacting risk for ASD in multiplex families as compared to simplex families. We note that multiplex/simplex status was indicated by either enrollment or self-report in a questionnaire and may be underestimated due to missing survey data.

As the prevalence of ASD is higher in males than in females (OR = 4.20) (63), and previous studies have reported that females with ASD have a higher burden of *de novo* variants (9,64–66), we also investigated the potential contribution of common variants to the female protective effect by comparing PRS between sexes. We did not find any evidence that ASD common variant risk burden differs in females compared to males (Figure 2C).

A previous study hypothesized that a new mutation in a mother, who is less susceptible to developing autism because of the female protective effect, may be more likely to transmit risk factors to their children with ASD (67). We, therefore, examined the over-transmission of common variant risk for ASD from mother to offspring. We found no evidence of the over-transmission of common variant risk burden from either mothers or fathers to their affected children (Figure 2D).

### Contribution of cortical development to risk for ASD

Previous studies suggest an important role of brain development in ASD (14,68). To characterize tissue types relevant to risk for ASD, we next evaluated heritability enrichment within active enhancer or promoter regions in different tissues (69) (Figure 3A, Table S4). Significant enrichment of heritability was observed in regulatory elements of brain germinal matrix as well as primary cultured neurospheres from the fetal cortex (FDR = 0.004 and 0.015, respectively), suggesting that disruption of gene regulation in these tissues increases the risk for ASD. We further examined SNP heritability in the developing cortex using differentially accessible peaks between the neuron-enriched cortical plate and the progenitor-enriched germinal zone (70) (Figure 3B). We found significant enrichment in peaks more accessible in the germinal zone (FDR = 0.008), but not in the cortical plate, providing further evidence for genetically mediated alterations in cortical development playing a crucial role in ASD etiology.

**Figure 3.**
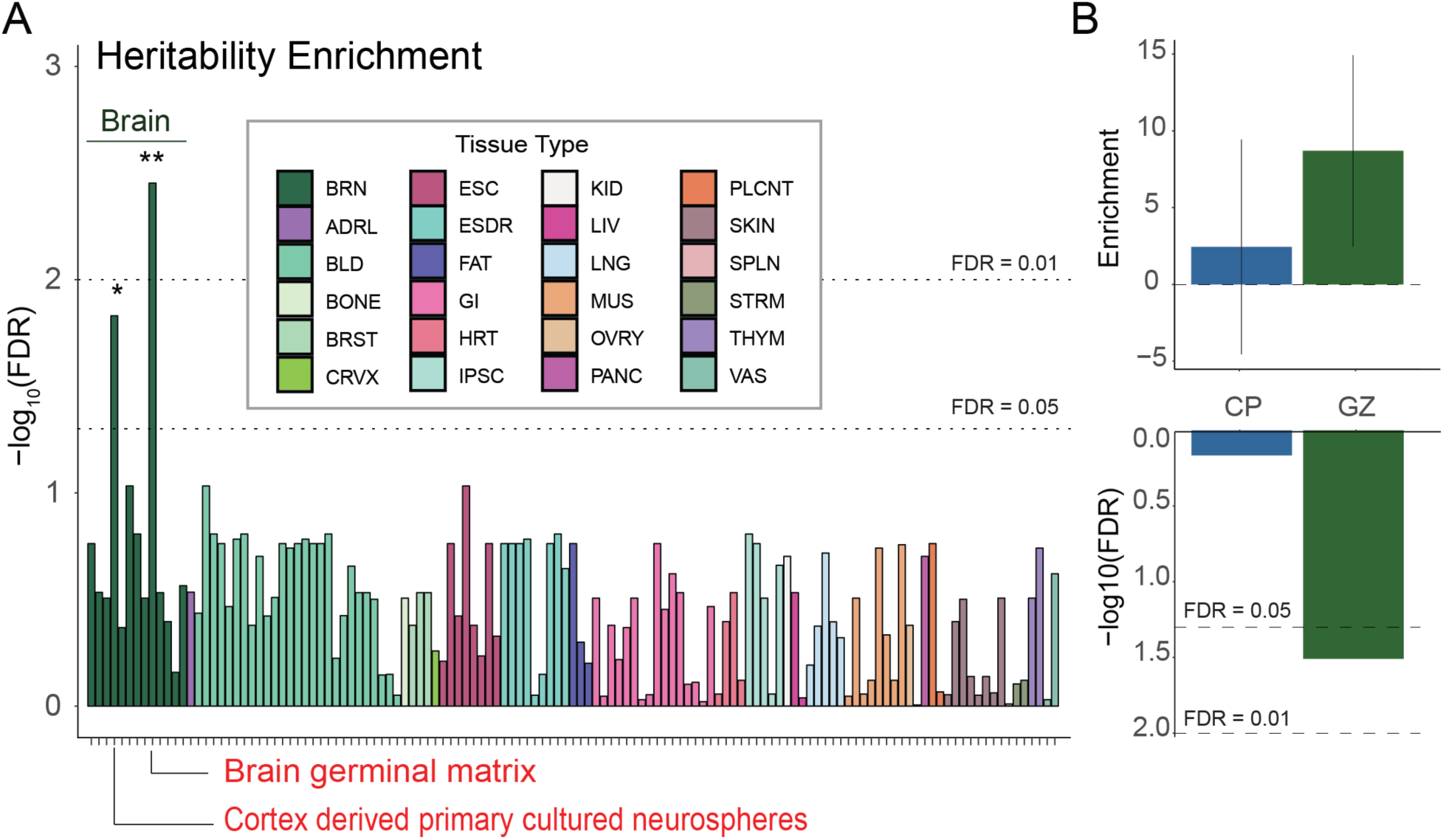
Partitioned heritability enrichment of tissues implicates cortical development in ASD risk. **(A)** Heritability enrichment in active enhancer and promoter regions present in different tissues shows the critical role of developing the brain in ASD etiology. **(B)** Heritability enrichment in differential chromatin accessibility from the developing fetal cortical wall. (upper) The x-axis represents tissue types and the y-axis indicates heritability enrichment. The error bar shows a 95% confidence interval. (lower) The x-axis represents tissue types and the y-axis indicates statistical significance as -log10(FDR). BRN: Brain, ADRL: Adrenal, BLD: Blood, BRST: Breast, CRVX: Cervix, ESDR: ESC_derived, GI: GI_duodenum, GI_colon, GI_rectum, GI_stomach, GI_intestine, GI_colon, GI_rectum, GI_duodenum and GI_esophagus, HRT: Heart, KID: Kideney, LIV: Liver, LNG: Lung, MUS: Muscle, OVRY: Ovary, PANC: Pancreas, PLCNT: Placenta, SPLN: Spleen, STRM: Stromal connective, THYM: Thymus, VAS: Vascular, CP: Peaks more accessible in cortical plate, GZ: Peaks more accessible in germinal zone. * FDR < 0.05, ** FDR < 0.01.

### H-MAGMA identified genes and pathways impacting risk for ASD

To identify genes associated with risk for ASD from meta-analysis (EUR only), we applied Hi-C coupled MAGMA (H-MAGMA) (31), which aggregates SNP-level P-values into a gene-level association statistic with an additional assignment of non-coding SNPs to their chromatin-interacting target genes generated from fetal brain Hi-C (46) (Figure 4A). We identified 567 genes associated with ASD (FDR < 0.1), including 263 protein coding genes (Figure 4B, Table S5). Five genes implicated from common variant evidence (*KMT2E, RAI1, BCL11A, FOXP1*, and *FOXP2*) also harbored an excess of rare variants associated with ASD (11). This overlap between rare and common ASD risk variants was more than expected by chance (hypergeometric *P* = 0.01; Figure 4C), corroborating the previous result that common and rare variation converges on the same genes and pathways (31,71). We also found that 14 H-MAGMA genes were also differentially expressed in the post-mortem cortex between individuals with ASD and neurotypical controls (up-regulated in ASD: *NFKB2, BTG1, RASGEF1B, TXNL4B, IFI16, WDR73* and *C2CD4A*; down-regulated in ASD: *PAFAH1B1, SEMA3G, DDHD2, GTDC1 ASH2L, USP19* and *ARIH2*; FDR < 0.05) (72) (Figure 4D). Rank-based gene ontology enrichment analysis (73) suggested that ASD risk genes were enriched in 188 terms including telencephalon development and regulation of synapse organization (Figure 4E, Supplementary Table S7).

**Figure 4.**
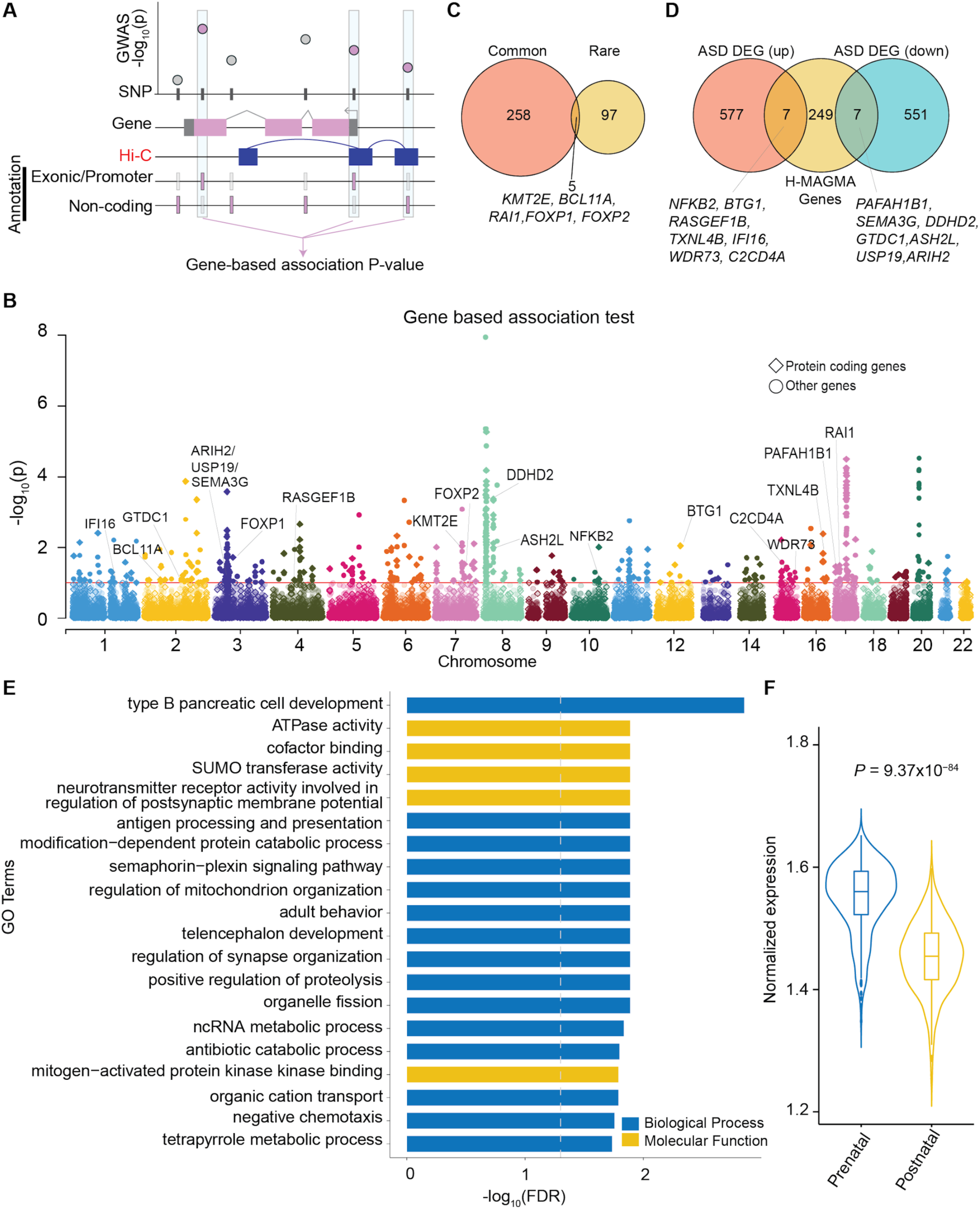
H-MAGMA identified 263 protein-coding genes linked to ASD. **(A)** Schematic diagram of H-MAGMA. SNP based association P-values were aggregated to gene-based P-values using positional information as well as chromatin interaction in the fetal brain. **(B)** Gene-based association result from H-MAGMA. The x-axis indicates the start position of genes (hg19). **(C)** Overlap of ASD risk genes harboring common and rare variants (11). **(D)** Overlapped genes with differential expression from post-mortem brains in individuals with ASD patients and neurotypical controls (72). **(E)** Gene ontologies enriched for ASD linked genes (top 20). **(F)** Developmental expression pattern of ASD risk genes (74).

Since heritability enrichment analyses suggested genetically mediated impacts on cortical development contribute to ASD risk (Figure 3), we explored whether the expression level of ASD risk genes from H-MAGMA is different between prenatal and postnatal cortex. As shown previously (14,31), we found ASD risk genes exhibited higher expression in the prenatal cortex as compared to the postnatal cortex (*P* = 2.77×10^−62^) (Figure 4F) (74). In particular, the expression level of ASD risk genes was highest between 20 and 30 post-conception weeks (Supplementary Figure S9). Taken together, our results demonstrate common risk variants for ASD play an important role in the developing cortex.

### Genetic correlation between ASD and 12 brain and behavioral phenotypes

Both epidemiological studies and genetic studies suggested the phenotypic comorbidity (75–78) or genetic correlation (14,79) of ASD with various brain and behavioral phenotypes. Thus, we evaluated the pleiotropic effect of ASD risk SNPs with twelve other brain and behavioral phenotypes (47,54,57,80–87) (Figure 5, Supplementary Table S8). We observed a novel genetic correlation between ASD and cigarettes per day (*r_g_* = 0.16, *P* = 8.80×10^−5^), indicating a partially shared genetic basis between risk for ASD and addictive smoking behavior. We also replicated positive genetic correlations previously detected for seven phenotypes (FDR < 0.05) (14), providing further support for a shared genetic basis of multiple neuropsychiatric disorders (79,88).

**Figure 5.**
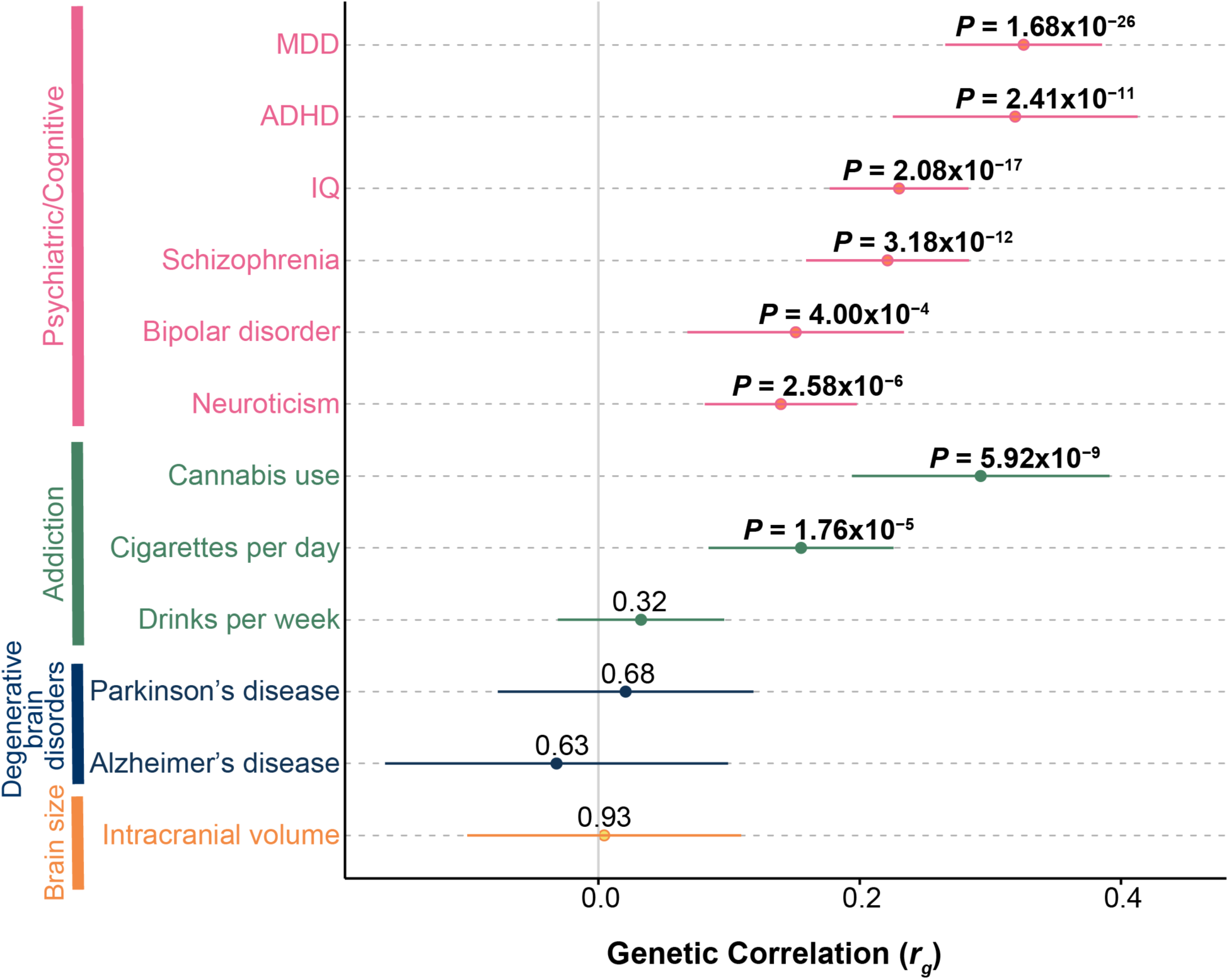
Genetic correlation of ASD against twelve brain and behavioral phenotypes. The x-axis represents an estimate of the genetic correlation (*r_g_*). Error bars represent the 95% confidence interval. P-values at FDR < 0.05 are shown in bold. MDD: Major depressive disorder, ADHD: Attention-Deficit/Hyperactivity Disorder.

### Functional validation to fine-map causal variants and prioritize genes

Interestingly, the novel locus identified by the SPARK full dataset (rs60527016 at chr8:38.19M - chr8:38.45M, Figure 1A, 6A) was also identified as a pleiotropic locus in a recent cross-disorder meta-analysis on eight psychiatric disorders (79). This locus was not only associated with ASD but also with schizophrenia, bipolar disorder and obsessive-compulsive disorder (OCD), suggesting that understanding the regulatory mechanism at this locus may reveal the basis for pleiotropic effects across psychiatric disorders. The index SNP (rs60527016) was located within a 300 kb LD block (*r*^*2*^ > 0.5 in SPARK full dataset) that contains seven genes (Figure 6A). To prioritize causal variants within this locus, we performed a massively parallel reporter assay (MPRA) (23,24) on 98 credible SNPs in this region in HEK 293 cells. MPRA measures barcoded transcriptional activity driven by each allele in a high-throughput fashion (Supplementary Figure S13). Surprisingly, SNP rs7001340 exhibited the strongest allelic difference in barcoded expression (*P* = 1.51×10^−24^) even though it is 37 kb away from the GWAS index SNP (*r*^*2*^ = 0.85 with the index SNP in SPARK full dataset) (Figure 6A, B, Supplementary Table S9), demonstrating the regulatory potential of this SNP and suggesting its causal role in psychiatric disorders, including ASD. While MPRA was performed in HEK cells, the SNP was located in a regulatory element with higher chromatin accessibility in human neural progenitors compared with postmitotic neurons (Figure 6A) (unpublished data from Stein lab), indicating its regulatory potential in the developing brain. The risk allele (T) at this SNP was associated with downregulation of barcoded expression in MPRA (Figure 6B), and was predicted to disrupt two transcription factor binding motifs (89) (TBX1 and SMARCC1) (Supplementary Figure S14), providing a possible mechanism of action of this variant. We next investigated potential target genes impacted by regulatory changes at this SNP by using eQTL data from fetal (33) and adult brain tissues (32). Expression levels of three eGenes were significantly associated with this SNP (*DDHD2* from the fetal brain and *DDHD2, LSM1, LETM2* from the adult brain) (Figure 6A). Of these three genes, two genes (*DDHD2, LETM2*) showed the direction of the effect expected from the MPRA result (risk allele downregulates the eGene) (Supplementary Figure S15). It is of note that *DDHD2* was identified in both fetal and adult brain eQTL datasets (beta = -0.080, *P* = 2.212×10^−13^; beta = -0.177, *P* = 1.38×10^−20^, respectively; Figure 6C, D). Notably, *DDHD2* was also significantly downregulated in the post-mortem cortex of individuals with autism (logFC = -0.28, FDR = 0.013), providing an added layer of evidence supporting its role in ASD risk (72). *DDHD2* also was identified by H-MAGMA (Figure 4F), and a copy number variation (CNV) containing *DDHD2* (deletions) was found in proband-sibling pairs with discordant social-behavior phenotypes (90). Collectively, by integrating existing multi-level functional genomic resources and an experimental fine-mapping approach using MPRA, we suggest *DDHD2* as a strong candidate gene impacting risk for ASD.

**Figure 6.**
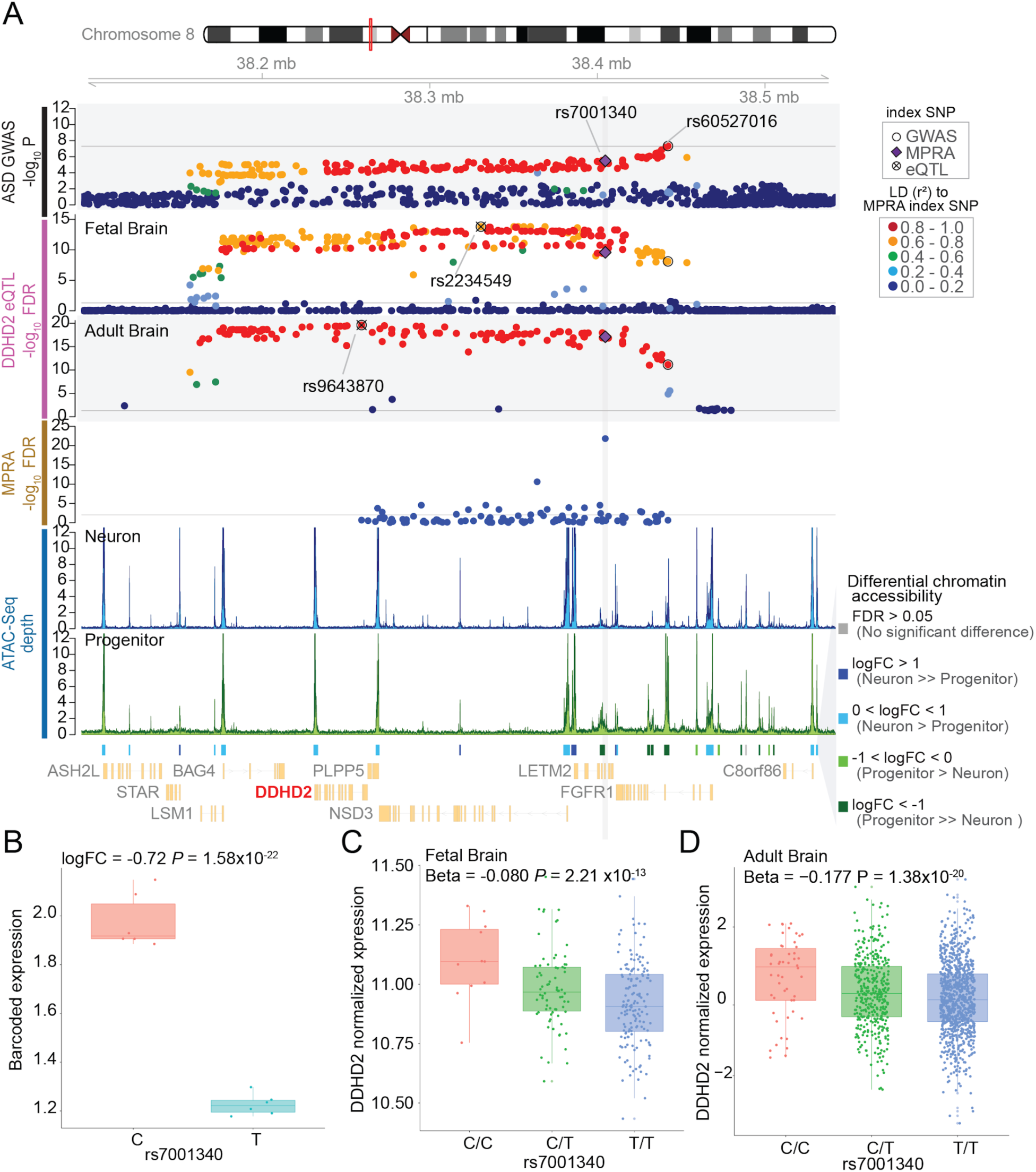
Identification of putative causal variant and gene impacting risk for ASD. **(A)** Annotated locus plot near rs60527016 ASD risk index variant, from top panel to bottom, ASD associations within SPARK full dataset (n = 6,222 case-pseudocontrol pairs), eQTL for DDHD2 in fetal brains (n = 235) and adult brain (n = 1,387), MPRA expression (n=6), ATAC-seq averaged depth in neuron (n = 61) and progenitor (n = 73). Differential open chromatin accessibility peaks from ATAC-seq, and gene model (NCBI Refseq). LD was calculated to rs7001340 within SPARK parents of cases, fetal brain donors, or 1KG EUR and colored accordingly. **(B)** The barcoded expression level of mRNA based from each allele at rs7001340 from the MPRA experiment. **(C)** The expression level of DDHD2 by rs7001340 genotypes in the fetal brain. **(D)** The expression level of DDHD2 by rs7001340 genotypes in adult brain. Individuals with allele dosage (0-0.1 as C/C, 0.9-1.1 as C/T, 1.9-2.0 as T/T) are shown. For (**B**) to (**D**), ASD risk allele for rs7001340 is T and protective allele is C.

## Discussion

In this study, we increased sample sizes for ASD GWAS to N_case(max)_ = 24,063, N_control(max)_ = 34,191 and identified five loci associated with risk for ASD (four from European only meta-analysis, one locus from SPARK project alone), including two new loci (marked by index SNPs rs60527016 and rs112436750). These loci have pleiotropic effects on multiple psychiatric disorders including schizophrenia (for rs60527016 and rs112436750), bipolar disorder, and OCD (for rs60527016).

Using a PRS derived from a previous study (14), we found enrichment of risk variants in SPARK cases, indicating the contribution of common genetic risk factors to ASD is consistent across cohorts. However, despite several hypotheses that rare variants associated with risk for ASD are enriched in certain subgroups of individuals with ASD, such as in females compared to males (female protective effect) (9,64–66,91), multiplex families compared to simplex families (12,61,62), or maternal alleles compared to paternal alleles (10,67), we do not find evidence to support the increased burden of common risk variants within those subgroups. This result indicates potential rare and common variant differences in contribution to subgroup risk for ASD. However, given the small sample size of specific subgroups (*N* = 835 in female whereas *N* = 3,262 in male, *N* = 14 for families with multiple affected children vs *N* = 3,618 with one affected children), our study may have limited power to identify the differences among subgroups. Thus, a larger sample size would be warranted to compare the difference in the role of common variants within these categories. Identifying locations in the genome associated with risk for ASD does not in itself lead to insights into what tissues or developmental time points are crucial for the etiology of ASD. Here, by integrating SNP association statistics with existing annotations of regulatory elements active during specific developmental time periods or within specific brain regions, we found an excess of common genetic risk for ASD in the fetal brain regulatory elements (brain germinal matrix and primary cultured neurospheres from the fetal cortex), and progenitor enriched germinal zone of the developing cortex, confirming previous findings that alterations of gene regulation in the prenatal cortex play a key role in ASD etiology (14).

To further understand the specific genes leading to risk for ASD, we applied a recently developed platform, H-MAGMA (31) and identified 263 putative candidate protein-coding risk genes. H-MAGMA genes are highly expressed in the prenatal brain, similar to the enrichment of ASD risk genes with rare variations during neurodevelopment (92). This result suggests potential molecular convergence regardless of classes of mutation, which is supported by five genes (previously identified *KMT2E* and newly identified *RAI1, BCL11A, FOXP1*, and *FOXP2*) that are affected by both rare and common variation.

Since identification of a GWS locus does not elucidate the causal variant(s), we performed MPRA and identified a potential causal SNP (rs7001340) at a novel ASD locus discovered in the SPARK sample. Interestingly, the individual variant with the strongest regulatory effect (rs7001340; *r*^*2*^= 0.85 with the index SNP in SPARK full dataset) was different from the SNP with the strongest association with ASD (rs60527016), highlighting the importance of experimental validation in identifying causal variants. It should be noted that the regulatory effects of these variants were assessed in non-neural (HEK) cells, so further validation of these effects in ASD-relevant cell types would provide increased confidence in declaring this SNP as causal. The experimentally validated regulatory SNP (rs7001340) is in the intron of *LETM2*, and is also an eQTL for *LETM2, LSM1* (247 kb away) and *DDHD2* (173 kb away), indicating that the SNP functions as a distal regulatory element. The risk allele (T) was associated with decreased expression of barcoded transcripts in the MPRA and downregulation of *DDHD2* from eQTL in both fetal and adult brains, implying a consistent direction of the allelic effects on gene regulation. The risk allele showed the same direction of effect for *LETM2* in adult brain tissue, but was not significantly associated in fetal brain tissue (P-values = 0.33). Intriguingly, *DDHD2* was also downregulated in the cortex from individuals with ASD compared to neurotypical controls (72), providing an additional level of support for this gene as a risk factor for ASD. *DDHD2* (DDHD domain-containing protein 2), also known as KIAA0725p, encodes a phospholipase and is localized in the Golgi (93). *DDHD2* plays a role in the efficient regulation of membrane trafficking between the Golgi and cytosol (93) and is highly expressed in the brain (94–96). Mutations in *DDHD2* have been previously found in individuals with spastic paraplegia type 54 (SPG) (96–98). *Ddhd2* null mice exhibited motor and cognitive impairments (99), which are frequent comorbidities of ASD (100). We, therefore, conclude *DDHD2* is a strong candidate risk gene for ASD through multiple lines of evidence.

There is still a large amount of common variant heritability not explained in this study indicating that further increases in sample size will be necessary to explain the common inherited component of ASD risk. While the combination of TOPMed imputation and the case-pseudocontrol study model enabled us to include individuals from multiple ancestries, the case-pseudocontrol model is lower powered compared to case-unscreened control models because a pseudocontrol might have greater liability for ASD than the average individual in the population (101). In addition, the case-pseudocontrol model cannot incorporate duos or singletons due to the lack of parental genotype information, which resulted in over 2,000 individuals with ASD with genotyping information in the SPARK project not being included in our analysis. Future studies could potentially increase power by including all cases available in SPARK and using unscreened population matched controls (102). Secondly, subsequent analyses including PRS, LDSC regression, and H-MAGMA were limited to individuals from European ancestries only, because most resources and software are designed to be used only within one population, generally European ancestry (103). Including other ancestries for these analyses will be able to uncover risk factors shared or specific to existing human populations.

In summary, ASD GWAS in the SPARK dataset and meta-analysis with previous GWAS identified two new susceptibility loci. By integrating existing multi-level functional genomic resources and experimental tools such as MPRA and eQTL, we highlight *DDHD2* as a novel high confidence ASD risk gene impacted by a distal common variant within a regulatory element present in neural progenitors of the developing cortex. This strategy can be broadly applied to common variant risk loci of multiple neuropsychiatric disorders to identify causal variant(s), regulatory regions, cell-types, and genes whose misregulation leads to risk for neuropsychiatric disorders.

## Data Availability

Approved researchers can obtain the SPARK population dataset described in this study (more details available at https://www.sfari.org/resource/spark/) by applying at https://base.sfari.org.
TOPMed study investigators contributed data to the reference panel, which can be accessed through the Michigan Imputation Server; see https://imputationserver.sph.umich.edu.
Summary statistics from the GWAS conducted here will be available upon publication.

## Acknowledgments

This work was supported by a grant from the Simon Foundation (SFARI[Award #: 605259], H.W and J.L.S), NIMH (R00MH113823 and DP2MH122403 to H.W. and R01MH118349, R01MH120125, R01MH121433 to J.L.S), NIGMS (DP2GM114829 to S.K.) and the NARSAD Young Investigator Award (H.W.). LMR was supported by T32 HL129982. We are grateful to all of the families in SPARK, the SPARK clinical sites and SPARK staff. We appreciate obtaining access to genetic and phenotypic data on SFARI Base. Approved researchers can obtain the SPARK population dataset described in this study (more details available at https://www.sfari.org/resource/spark/) by applying at https://base.sfari.org.

Trans-Omics in Precision Medicine (TOPMed) program imputation panel (version Freeze5) supported by the National Heart, Lung and Blood Institute (NHLBI); see www.nhlbiwgs.org. TOPMed study investigators contributed data to the reference panel, which can be accessed through the Michigan Imputation Server; see https://imputationserver.sph.umich.edu. The panel was constructed and implemented by the TOPMed Informatics Research Center at the University of Michigan (3R01HL-117626-02S1; contract HHSN268201800002I). The TOPMed Data Coordinating Center (3R01HL-120393-02S1; contract HHSN268201800001I) provided additional data management, sample identity checks, and overall program coordination and support. We gratefully acknowledge the studies and participants who provided biological samples and data for TOPMed. Summary statistics from the GWAS conducted here will be available upon publication. Code is available at https://github.com/thewonlab/GWAS_ASD_SPARK.

